# Group Testing Performance Evaluation for SARS-COV-2 Massive Scale Screening and Testing

**DOI:** 10.1101/2020.05.02.20080390

**Authors:** Ozkan Ufuk Nalbantoglu

**Affiliations:** Department of Computer Engineering and Genome and Stem Cell Center, Erciyes University, Kayseri, Turkey

## Abstract

The capacity of current molecular testing convention does not allow high-throughput and community level scans of COVID-19 infections. The diameter in current paradigm of shallow tracing is unlikely to reach the silent clusters that might be as important as the symptomatic cases in the spread of the disease. Group testing is a feasible and promising approach when the resources are scarce and when a relatively low prevalence regime is observed on the population. We employed group testing with a sparse random pooling scheme and conventional group test decoding algorithms both for exact and inexact recovery. Our simulations showed that significant reduction in per case test numbers (or expansion in total test numbers preserving the number of actual tests conducted) for very sparse prevalence regimes is available. Currently proposed COVID-19 group testing schemes offer a gain up to 10X scale-up. There is a good probability that the required scale up to achieve massive scale testing might be greater in certain scenarios. We investigated if further improvement is available, especially in sparse prevalence occurrence where outbreaks are needed to be avoided by population scans. Our simulations show that sparse random pooling can provide improved efficiency gains compared to row-column group testing or Reed-Solomon error correcting codes. Therefore, we propose that special designs for different scenarios could be available and it is possible to scale up testing capabilities significantly.

## 1 Introduction

The first half of the year 2020 has been the midst of a first wave of COVID-19 pandemic. It is expected that, with the strict social distancing applications, the spread of SARS-CoV-2 is going to be stabilized or sustained at several geographies [1]. However, second wave epidemics and resurgences are highly likely to occur [2]. Moreover, different geographies, due to segregation by administrative borders, asynchronously experience epidemic growths. While social distancing and quarantine measures have been the primary factor to contain SARS-CoV-2, widespread testing and aggressive contact tracing are observed to be effective to isolate the spreaders from vulnerable populations. According to this, widespread scanning of populations or subpopulations is crucial to locate and isolate epidemic clusters, especially by detecting asymptomatic carriers. Excessive numbers of asymptomatic carriers appear to be especially important as they might be the most important factor in the difficulty of lowering the reproductive numbers. An unbiased estimation of the ratio of asymptomatic or presymptomatic spreaders might be difficult to assess. Yet, statistics from small or medium sized cohorts and case studies indicate that they might be as abundant as 10%-50% [3] [4]. These ratios possess special importance regarding the case studies observing that asymptomatic/presymptomatic carriers are likely to infect their contacts [5] [6].

Currently, PCR-based molecular testing primarily serves as the standard diagnostics in the majority of health systems. Since the infrastructure and financial limitations dictate the allocation of resources for higher priority cases, the capacity of current molecular testing convention does not allow high-throughput and community level scans. Therefore, the focus of testing is on hospitalized symptomatic cases and subsets of their contacts reached via tracing. The diameter in current paradigm of shallow tracing is unlikely to reach the silent clusters that might be as important as the symptomatic cases in the spread of the disease.

Group testing is a feasible approach when the resources are scarce and when a relatively low prevalence regime is observed on the population [7]. It allows scanning large populations by pooling samples and conducting orders of magnitude lower number of tests, while being able to locate the positive cases. Starting from its initial practical use in US Army in the 1940s [8], screening new recruits for syphilis, group testing has been popularly employed in several different fields where pooling/mixing and testing was available to detect the defective elements. In general, group testing algorithms seek for 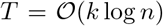 measurements in order to correctly detect *k* positive cases in *n* testing samples. Practical algorithms are shown to achieve 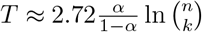 while *k* scales sublinearly as *k* = *θ*(*n^α^*) [9]. With an assumption that early phases of epidemic waves or screening of asymptotic carriers would form populations of relatively low prevalence, conventional group testing schemes can potentially provide significant gains in testing capability.

Complying with the idea of employing group testing in SARS-CoV-2 screening, Sinnott-Armstrong et al. [10] recently proposed an adaptive scheme, pooling rows and columns of PCR well-plates, and they showed that up to 10X efficiency gain could be achieved in 0.1%-5% prevalence band. While this is a very practical and a simple scheme eliminating the requirements of robotic pipetting needs and reducing the pipetting errors, more sophisticated pooling schemes that can improve the efficiency gains are available. Indeed, Shental et al. [11] used sampling 48 pools out of 384 well-plate by Reed-Solomon coding, and showed 8X efficiency gain around the band of 1% prevalence. These schemes offer very significant scale-ups in testing capacity, especially around mid-scale sparsity bands (i.e. 1%-5% prevalence). However, in potential scenarios of achieving wide-spread or community level scans with lower expected prevalence, further efficiency gains would be required. Therefore, designing group testing schemes aiming sparsity is beneficial to explore the practical capabilities.

We employed group testing with a sparse random pooling scheme and conventional group test decoding algorithms both for exact and inexact recovery. Our simulations showed that significant reduction in per case test numbers (or expansion in total test numbers preserving the number of actual tests conducted) for very sparse prevalence regimes is available.

## 2 Method

We designed a random-pooling based group testing scheme for different laboratory set-ups including 96-, 384-, and 1536-well-plate options. Two different approaches were considered i) two-pass adaptive testing for exact recovery. ii) single-pass non-adaptive testing for approximate recovery of test results. For both approaches, we use the same random pooling design.

### Random pooling

Random pooling procedure samples a pooling procedure from the space of m pools out of n samples by adding each sample exactly to k pools. The distribution to pools are selected without replicating the same pattern in order to be able to distinguish samples. Also, the sampled pooling matrices are intended to have low coherence, as low coherence improves the rate (i.e. efficiency gain in this setting) in sparse recovery problems. We achieve low coherence by Metropolis sampling algorithm [12] with a modification that acceptance is driven by coherence, rather than density.

**Algorithm 1:**
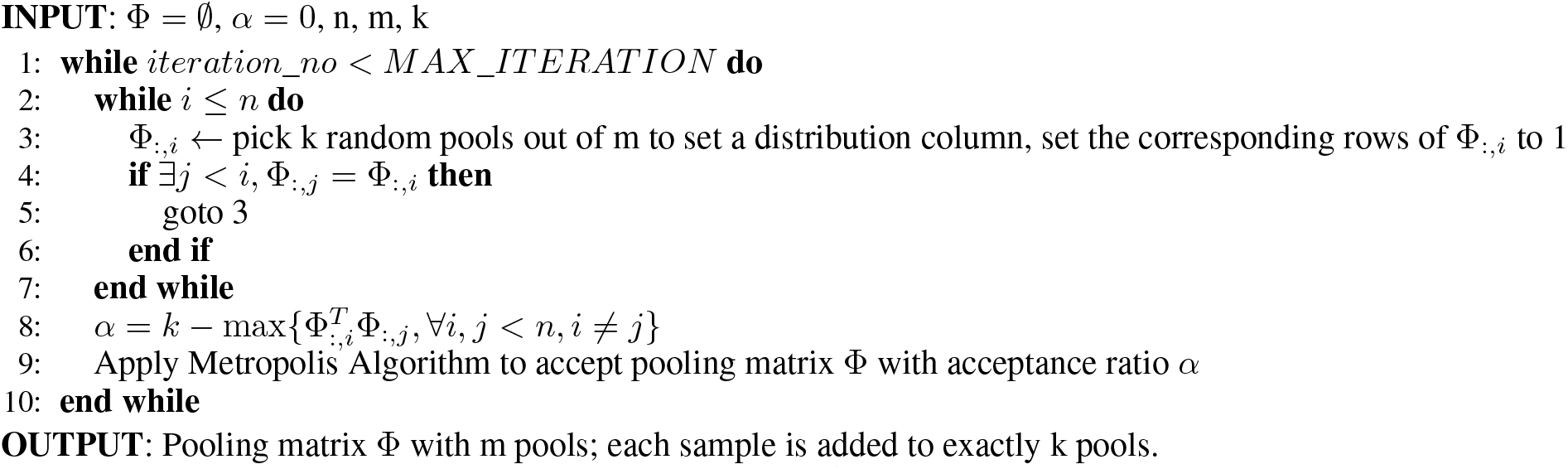
Random pooling

### Two-pass testing

This scheme concentrates on two rounds of testing. The first test round employs random pooling and testing of pooled samples. Samples that are confidently negative are eliminated using a possible positive detection algorithm. At this point, a subset of the samples are determined as possible positives for further testing, and no false negatives are eliminated. Testing the possible positives directly for eliminating the false positives concludes the testing process without false calls. Note that, perfect recovery in this sense assumes single tests are accurate.

We have employed the straightforward “Definite Defectives” algorithm [13] to detect the possible positives in the first round. Once the possible positives are subject to the second round, the remaining procedure includes conventional “one test-one sample” approach. The procedure is given as follows.

**Algorithm 2:**
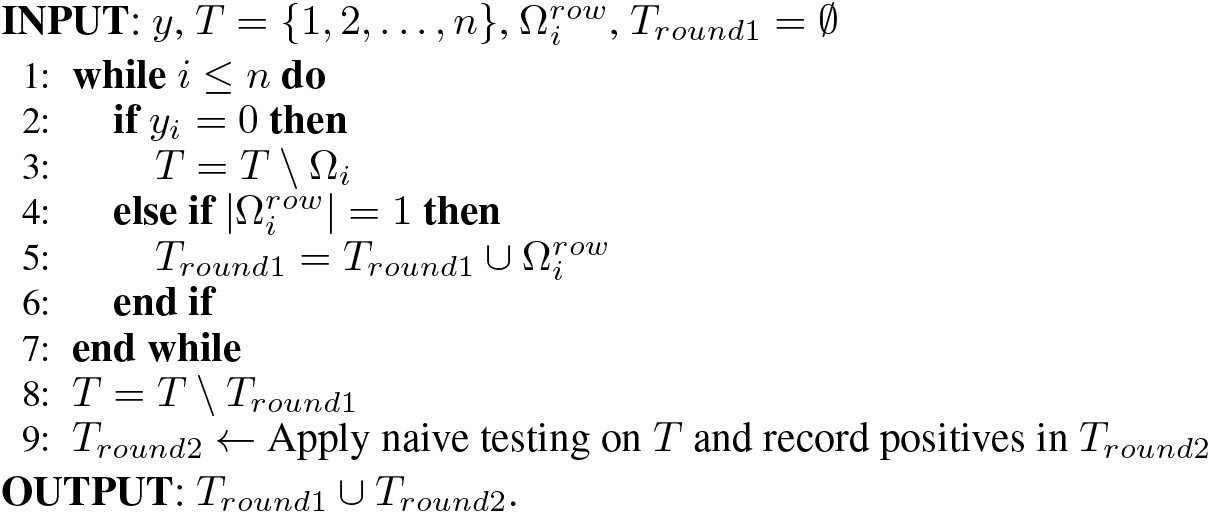
Two-pass testing (Definitive Defectives)

Here, we refer to the binary vector *y* as the pooled test results where 1s are the positive pool, and 0s are the negative pool results. 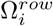 represents the support set of *i^th^* row in Φ pooling matrix determined by random pooling scheme, and *T* sets represent the set of positive candidates.

### Single pass testing

Single pass testing includes only the approximate decoding of pooled samples in one round. We considered Sequential Combinatorial orthogonal matching pursuit (COMP) algorithm [13] for decoding. According to this, among the set of possible defectives, the minimal set explaining the pooling test results is considered using a greedy forward addition. This set is the predicted positives for the entire test population. Unlike previous approach, single pass testing is vulnerable to false positives and negatives, whenever the pooling scheme is underpowered to explain the actual population uniquely.

**Algorithm 3:**
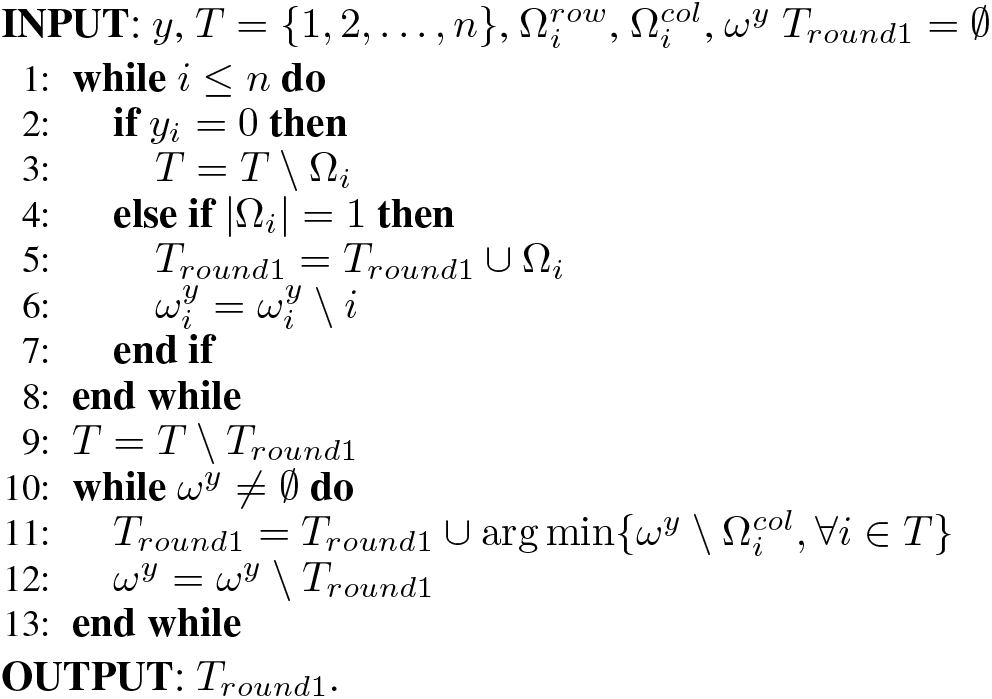
Single pass testing (COMP)

Here, similarly we refer to the binary vector *y* as the pooled test results. *ω^y^* represents the support set of *y*, 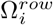 represents the support set of *i^th^* row in Φ, 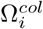 represents the support set of *i^th^* column in Φ, and *T* sets represent the set of positive candidates.

While the pooling procedure is stochastic, it is possible to design several different pooling schemes. As a laboratory adopts one, the decoding algorithm needs to know the pooling procedure to construct a mixing matrix and solve.

## 3 Results

In order to observe the performance of group testing under different regimes, simulations under Bernoulli sampling were repeated and reported. The prevalence range between 0.0005 and 0.2 was chosen as the simulation operating region. Efficiency gain, defined as the ratio of tested subjects to the number of actual tests performed in pools was used as the performance metric. Three pooling strategies, each corresponding to a different plate operation were considered: 96-well plate, 384-well plate, and 1536-well plate.

### i) Two-pass testing

In case of exact recovery of one sample-one test naive testing, which would fix the sensitivity and specificity of group testing approximately to that of PCR technical limitations, the two-pass procedure explained the methods, referred to as “Definitive defectives” was considered. For 96-well plate set up, 10 pools are considered for the first round group testing and each sample is represented exactly in three different pools. The preferred number of pools was 15 with each sample is distributed to exactly 4 pools for 384-well plate setting, also 20 pools with each sample being distributed to exactly 4 different pools was considered for 1536-well plate set up. The corresponding random pooling strategies construct sparse mixing matrices which is expected to result in good performance of decoding algorithms (i.e. detecting a small number of possible positives with ideal specificity). Consequently, greater efficiency gain values are expected compared to conventional grouping.

In order to observe how this pooling strategy scales for very sparse settings compared to previously proposed techniques, we considered the group testing scheme suggested by Sinnott-Armstrong et al. (which we refer to as “row-column grouping”) for the same setup. In all three pooling schemes, the proposed group testing resulted in more than two times higher efficiency in sparse prevalence regimes (Figure 1). Each observed value is the recorded mean of 1000 independent simulations. This came with the trade-off of underperforming in denser areas (0.2% to 0.1% regions).

**Figure 1:**
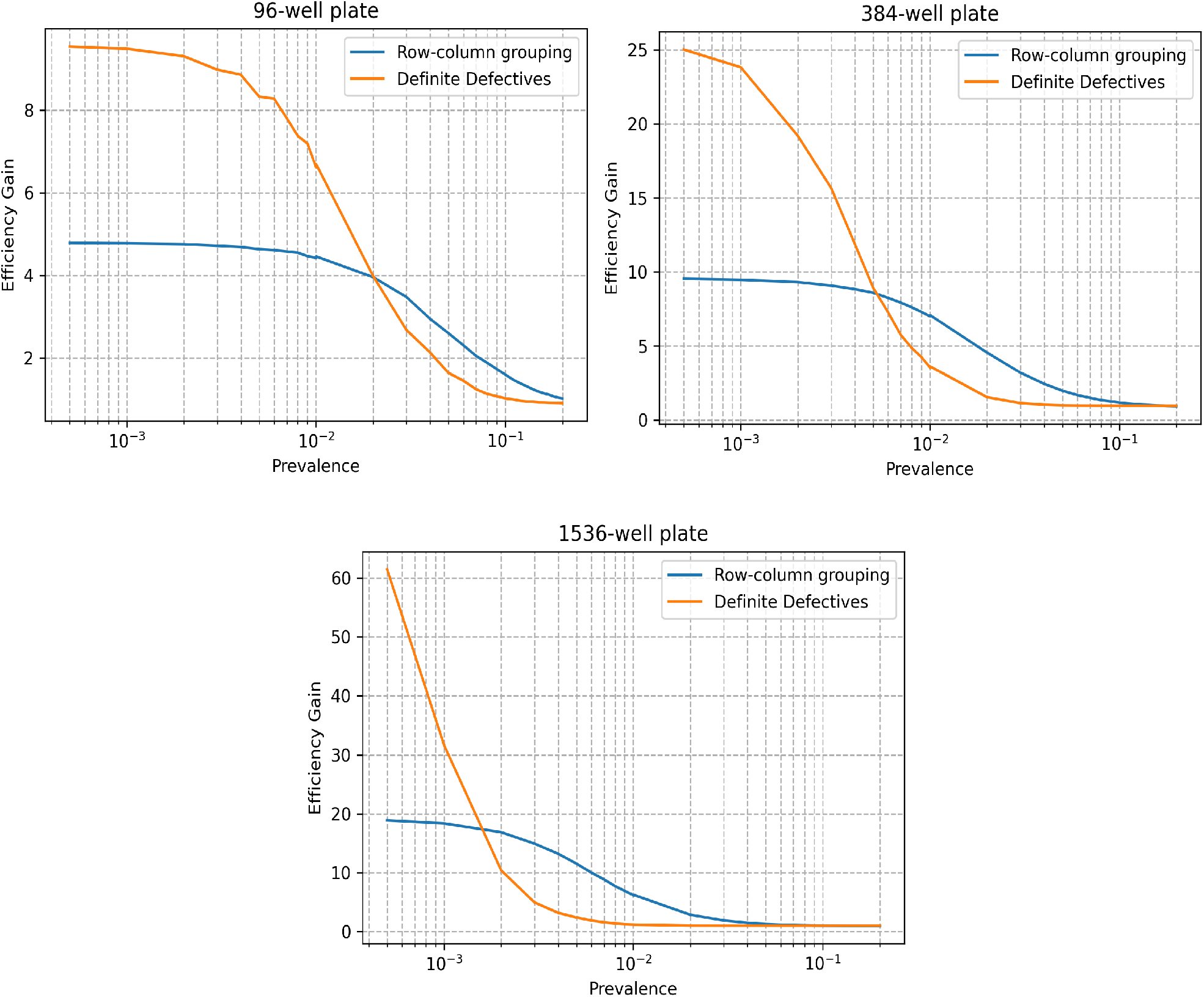
Efficiency gains of two-pass group testing schemes. Row-column testing defined by Sinnott-Armstrong et al. (row-column grouping) and the proposed random pooling (Definitive Defectives) were simulated for 96-, 384-, and 1536-well plate setups at a range of prevalence scenarios.

### ii) Single-pass testing

We have considered the scenarios which the preservation of naive test accuracy is not the primary concern. A widespread scanning concept might be focusing on determining the hot-spots within a population rather than diagnosing each individual for COVID-19. In such a case, test accuracy could be traded off in return of gains in testing capacity. Since single-pass group testing schemes are underdetermined systems analogous to compressed sensing, reduction in number of pools to be tested will translate to reduction in testing accuracy, which corresponds to the trade-off operation between efficiency gain and test accuracy. To observe this characteristics we conducted the simulations in similar prevalence ranges as the two-pass process. However, in this case random pools were decoded to sample test results in single pass using sCOMP decoding as described in the methods section. The accuracy increase was recorded with respect to increasing number of random pools at each prevalence instance. Starting from the same setting with two-pass testing pooling scheme, new pooling procedures were sampled at each pooling expansion. the parameter k, that corresponds to the contribution of a sample to the number of pools were incremented by one by the introduction of every 10 new pools.

As the performance metric, we chose sensitivity to reflect the operation success of group testing scheme. As expected, the specificity measures were close to 1 throughout the operational regions (supplementary figure 1). Figure 2 shows the efficiency gain operations, indicating operation at each sensitivity level as separate contours. Following similar characteristics with two-pass testing, pooling from larger well plates yields greater gains, especially for very sparse regimes. Typically, the trend of equal-sensitivity contours exhibit greater slopes as the prevalence gets smaller, indicating better efficiency gain boosts for lower sensitivity trade-off.

**Figure 2:**
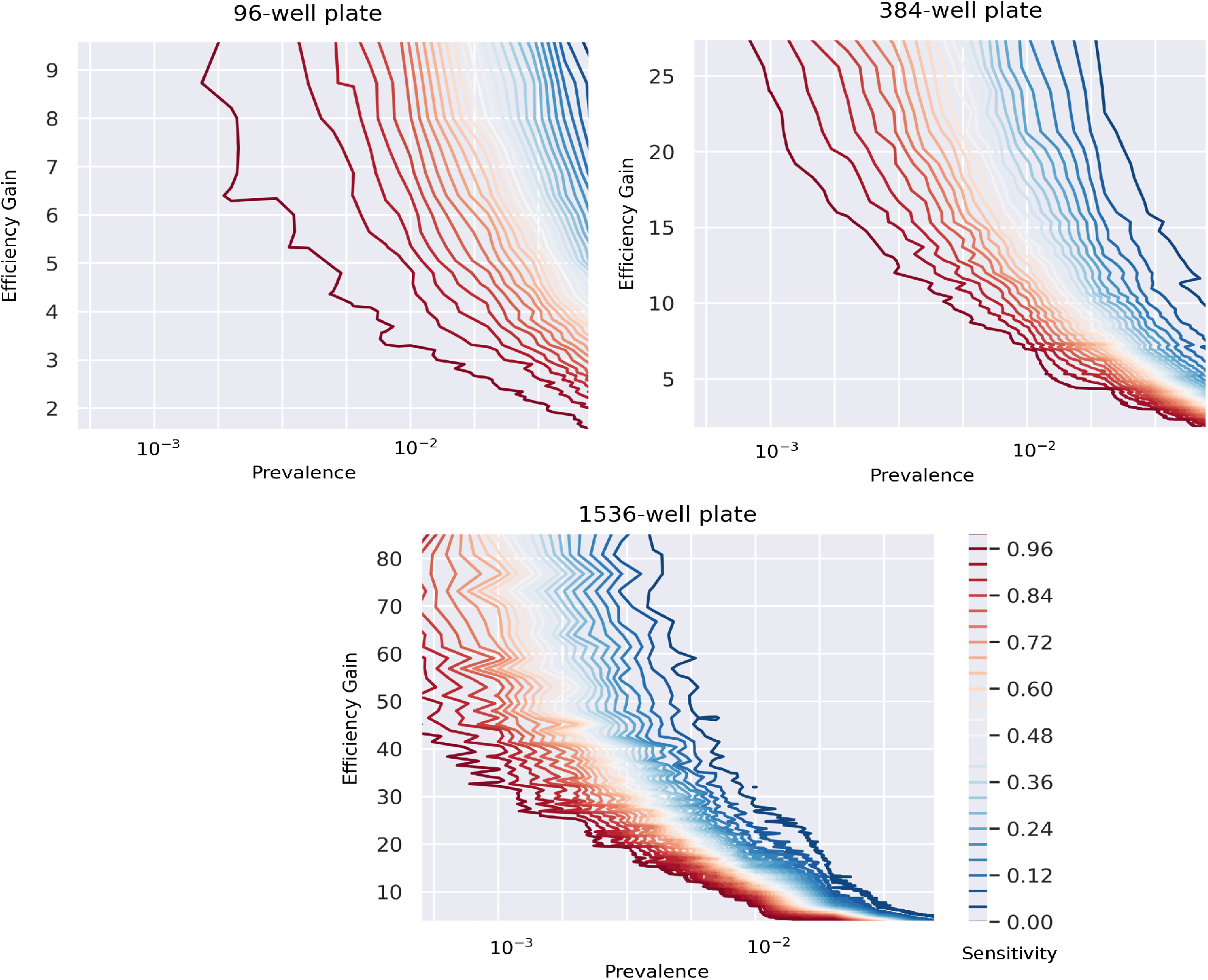
Efficiency gains of single-pass group testing. Decoding performed using sequential COMP for different number of pools were simulated for 96-, 384-, and 1536-well plate setups at a range of prevalence scenarios. As the number of pools reduces and the efficiency gain increases, the sensitivity of the test reduces. Equal-sensitivity contours are shown in different color codings.

### Single-pass vs. two-pass testing

Although single pass and two-pass schemes presented are not directly comparable since the former is a partial recovery method and the latter is an exact recovery method, it is interesting to compare their efficiency gains where single-pass testing performs close to exact recovery. According to our simulations, operation characteristics of single pass testing converges to row-column group testing as the prevalence increases, and it operates close to the proposed two-pass testing in low-prevalence regions (supplementary figure 2). It might be possible to perform single-pass testing with appropriate random pooling schemes even in high prevalence regimes.

### Single-pass testing vs. P-BEST testing

P-BEST testing was shown to perform accurately around the band 0.2%0.13% with 8X efficiency gain. While P-TEST incorporates a specific Reed-Solomon error-correcting code scheme for pooling, we ran simulations around this band with similar aspects (i.e. each sample contributes to exactly 6 pools in 384-well plate setting) to observe at what efficiency gain rates a perfect recovery is available. According to the averages over 3000 simulations, random pooling performs similarly around 1% prevalence (table 1). Note that this observation is not based on practical experiments neglecting any physical source of noise. In order to validate the results and draw a confident conclusion, further studies and wet-lab experiments are needed.

**Table 1:** Single-pass testing vs. P-BEST testing

| Prevalence | 0.26% | 0.52% | 0.78% | 1.04% | 1.3% |
| --- | --- | --- | --- | --- | --- |
| P-BEST | 8X | 8X | 8X | 8X | 8X |
| Single-pass | 16X | 10.66X | 8X | 8X | 6X |

## 4 Discussion

Conducting an effective test, trace isolate solution might be the key for containing SARS-CoV-2 and an elemental part of fighting against the current COVID-19 pandemic. Perhaps the most significant and challenging issue is performing massive-scale tests. Asymptomatic cases, being the possible silent culprits of undetected infection clusters, are off the radar of conventional testing procedures. In order to expand the radius of contact tracing to subpopulation scan level or performing community scans, given the current testing infrastructures and resources efficient group testing strategies are needed. Currently proposed COVID-19 group testing schemes offer a gain up to 10X scale-up. There is a good probability that the required scale up to achieve massive scale testing might be greater in certain scenarios. We investigated if further improvement is available, especially in sparse prevalence occurrence where outbreaks are needed to be avoided by population scans. Our simulations show that sparse random pooling can provide improved efficiency gains compared to row-column group testing or Reed-Solomon error correcting codes. Therefore, we propose that special designs for different scenarios could be available and it is possible to scale up testing capabilities significantly.

Our simulations indicate that the greatest efficiency gains promised by the proposed pooling schemes are achieved at very low prevalence rates. This observation leads to the fact that such a high-efficiency gain approach should be limited to certain testing scenarios. The proposed group testing, along with the other approaches reviewed previously, would not be helpful in the diagnostic use for symptomatic cases around outbreak peaks, for instance when the prevalence in a tested population is greater than 10%. On the other hand, a few cases at the early phases of an outbreak are worth to detect, as they might be the seeds for exponential growth of epidemic curves. Widespread testing for early detection of emerging clusters, therefore might be a powerful preventive approach. Our results claim that random pooling might be a preferable group testing strategy in similar scenarios. Another use case might be scanning for asymptomatic cases, separate from diagnostic testing on symptomatic individuals. This might be either as extending contact tracing to be able to test greater number of case contacts, or as a periodic scan of a population (e.g. healthcare workers and other risk groups). In case of community scans in which individual-level sensitivity can be sacrificed in return of spanning larger populations, single-pass group tests might also be useful. We have observed that for single-pass group testing, a small decrease in sensitivity could enable large efficiency gains at low prevalence groups. Such a high-throughput group testing on relatively large populations could be devised as a tool of locating infection hot spots.

Assume that contact tracing in a relatively low prevalence cluster is being conducted using naive testing. It might be useful to extend tracing by testing several fold more individuals but with the same number of tests, using group testing. The extended subpopulation may be very low prevalence, however detecting any asymptomatic case in it could be very valuable. We ran simulations to investigate what should be the initial prevalence for such an expansion opportunity, and how many folds of extension would be available in the proposed group testing scheme. We observed that up to 60X expansion is available around 2%-3% prevalence band (Supplementary figure 2). This result might be an implication that large scale contact tracing might be possible at early forming clusters.

It should be kept in mind that our experiments are simulations designed on two major assumptions. First, we assumed that pooling will not attenuate the detection efficiency of molecular detection. Yelin et al. [14] showed that pooling a single positive SARS-CoV-2 sample with tens of negative samples does not harm detectability of positive in care of TR-qPCR testing. The pooling size in our simulations does not exceed the experimented limits, so we believe proposed pooling schemes should act similarly in practical settings. Our second assumption is that the molecular tests are accurate enough to detect the samples without errors. Therefore, all simulations are performed for a noiseless scenario. While the actual picture might not comply with this assumption, the experiments ran with P-BEST approach indicate that imperfection of RT-qPCR tests does not have a negative effect on group testing. In fact, the encoding procedures of mixing matrices might even work as error correcting codes for low prevalence. However, further studies and wet-laboratory experiments should be conducted to support this claim.

Another aspect about group testing is that the efficiency gains mentioned in this and previous studies do not translate as linear gains in SARS-CoV-2 testing economy. Group testing only amplifies the number of tests, however previous steps of sampling, logistics and any sample preparation steps remain unscaled. Feasibility of pre-pooling operations is out of the scope where it might be another serious concern. Moreover, group testing might be subject to greater use of consumables (e.g. more pipetting), imposing the risk of mixing errors or the need of robotics systems.

Although current popular approach of COVID-19 testing RT-qPCR based molecular tests, group testing clearly is not limited to PCR based molecular techniques. Any testing scheme preserving positivity as well as lack of signals in the absence of positives while combining samples in pools can be trivially integrated to group testing approaches. IgC/IgM based serological tests or any prospective test strategies (e.g. CRISPR-Cas12 based testing [15]) should be considered for large scale testing/scanning with group testing under appropriate circumstances. It should be noted that the current concept of group testing relies on recovering the actual sample profile from binary outcomes of pooled tests. Testing schemes allowing quantitative outputs might be possible and further quantitative information might enable unprecedented efficiency gains. This opportunity can be hypothesized following the results of compressed sensing literature. For example, for the case of Sudocodes [16], at a prevalence of 0.1%, 1 Million subjects can be scanned by performing under 10.000 tests even with perfect recovery [36]. Of course, this requires very precise quantitative and unique measures for each test subject. It might be a promising research direction to investigate such testing options. For example, shallow shotgun sequencing allowing positive signals while pronouncing specific sequences belonging to each sample could be a candidate for ultra-high throughput group testing. Proposed group testing algorithms assume and simulate under the circumstances that each sampled instance is independent. On the contrary, we know that COVID-19 sampling is mainly conducted on populations with social contacts, violating the independence assumptions. Sophisticated decoding algorithms informed by metadata might perform superior, allowing greater efficiency gains, while complicating the overall testing procedure in practice. Nevertheless, we believe that even in primitive forms group testing is a promising direction for efficient allocation of testing resources that should be considered for practical use.

## Data Availability

All code is available, please contract the author at

## Supplement

### Accuracy of Single-Pass testing

Sequential COMP decoding employed for single pass decoding of group testing by random pooling operates on a greedy algorithm, selecting a suboptimal minimal subset satisfying the pool test results. Clearly, the solutions are not unique, and in case several linear combinations cover the same support set of pools, the decoding is vulnerable to errors. Lowering the number of random pools drives the system to be more underdetermined, expecting greater number of errors on average. This can be observed from our simulations. When we observe the empirical results, the specificity has never been experienced to be lower than 0.97, which is expected since the negative/positive distribution is unbalanced. This is also an indication that the decoding is not overexploring the positives. A more rigorous or liberal decoding scheme, allowing relaxations such as linear programming might be further investigated to observe if any improvement in decoding accuracy is achievable emprically.

### Single-pass vs. two-pass testing

Figure S2 provides the efficiency gain mean performance of the two proposed schemes. Roughly, the single pass scheme approximates the gains of random pooling and row-column pooling schemes at both sides of the prevalence spectrum. The single pass scheme appears to perform similar to two-pass complete recovery around near errorless operation (e.g. sensitivity >0.99). While this is practically preferable, as single-pass testing requires only one round of testing and it might be more efficient in time requirements, it might be more difficult to design a single-pass test. It should be noted that each efficiency gain requires different random pooling numbers for different prevalence regimes and a correct guess of prevalence should be predicted beforehand. In case prevalence distribution attains high variance, two-pass testing might be a safer approach.

**Figure S1:**
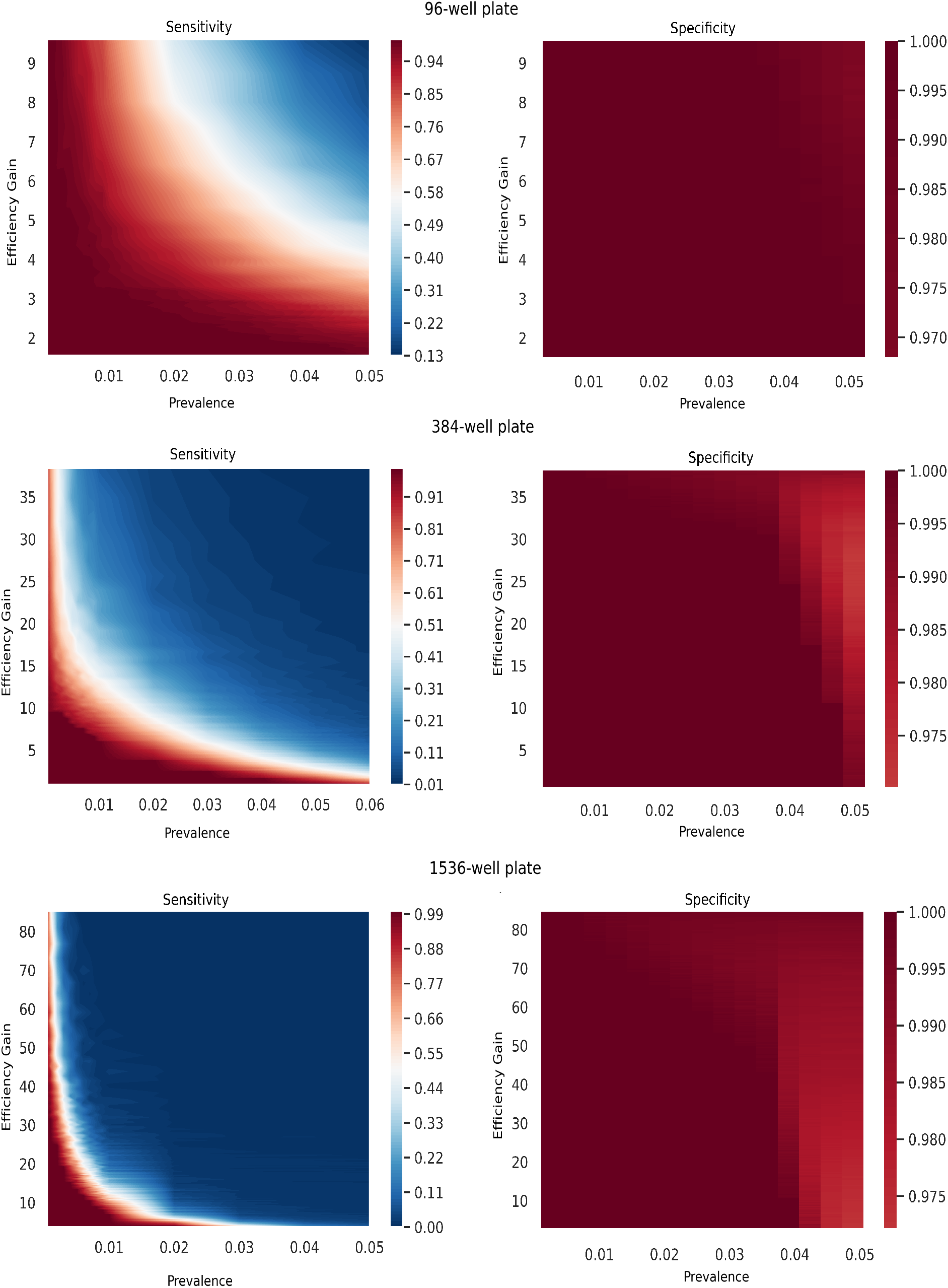
Sensitivity and Specificity of the proposed single-pass testing at different prevalences across different number of pools (i.e. reflecting as efficiency gains).

**Figure S2:**
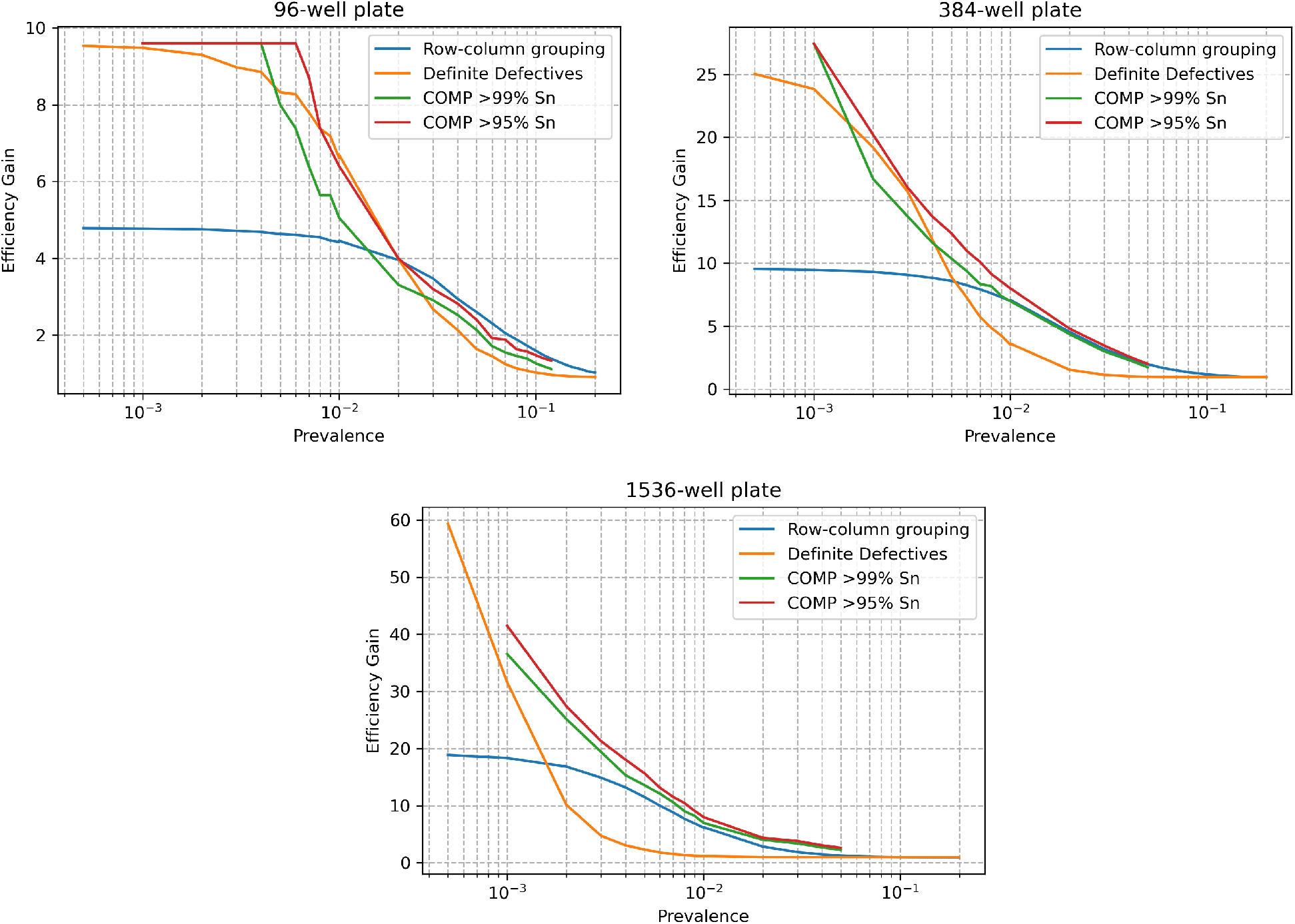
Efficiency gains of single-pass and two-pass schemes at different prevalences.

### Expansion rates

Figure S3 depicts the expansion rates, showing how many folds the testing capacity can be expanded to include presumably negative dominant scan population (e.g. extending the tracing to not only symptomatic but also to asymptomatic contacts) conducting the same number of tests. As expected, it would be possible to achieve larger expansions at low prevalences, especially with large plate setups. On the other hand, still room for a few-fold expansion seems to be achievable around relatively large prevalences such as 10%. This might be an indicator that group testing could be a feasible operation at large spectrum.

**Figure S3:**
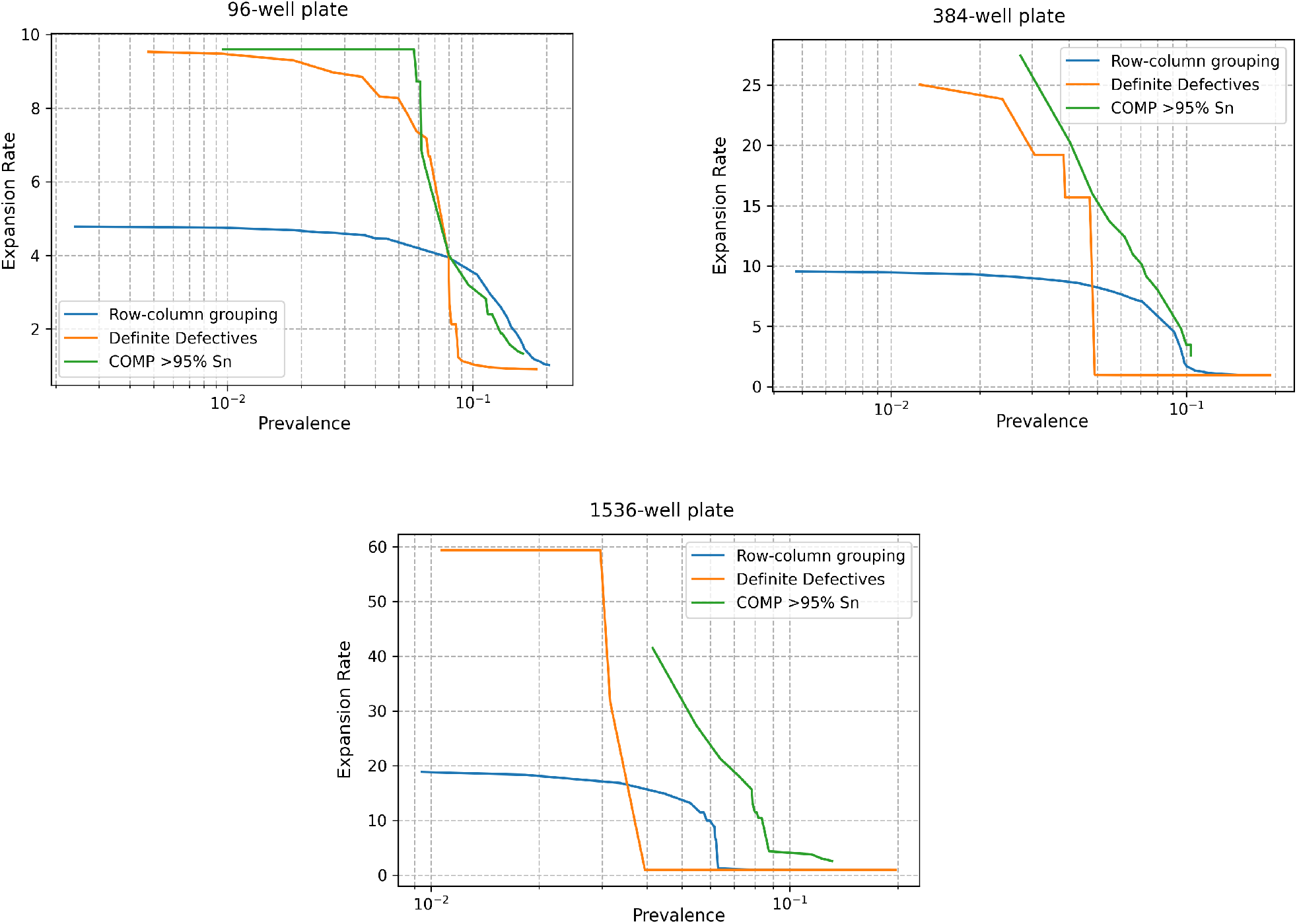
Expansion rates of single-pass and two-pass schemes at different prevalences.

